# Finding effective therapeutic approaches for motor rehabilitation after stroke: Insights from a thematic analysis

**DOI:** 10.1101/2021.08.01.21261366

**Authors:** Rajiv Ranganathan, Carson Doherty, Michael Gussert, Eva Kaplinski, Mary Koje, Chandramouli Krishnan

**Affiliations:** Department of Kinesiology, Michigan State University, East Lansing, MI, USA; Department of Mechanical Engineering, Michigan State University, East Lansing, MI, USA; Neuromuscular and Rehabilitation Robotics Laboratory (NeuRRo Lab), Department of Physical Medicine and Rehabilitation, University of Michigan, Ann Arbor, MI, USA

**Keywords:** rationale, ingredients, sample size, power, specificity, impairment

## Abstract

Despite tremendous advances in the treatment and management of stroke, restoring motor and functional outcomes after stroke continues to be a major clinical challenge. Given the wide range of approaches used in motor rehabilitation, several commentaries have highlighted the lack of a clear scientific premise for different interventions as one critical factor that has led to suboptimal study outcomes. To examine this issue in greater detail, we conducted a thematic analysis of randomized controlled trials in stroke rehabilitation over a 2-year period from 2019-2020. Our results revealed three primary findings: (i) most studies did not provide an explicit rationale for why the treatment would be expected to work, (ii) there was not a close correspondence between the active ingredients mentioned versus the active ingredients measured in the study, and (iii) multimodal approaches that involved more than one therapeutic approach tended to be combined in an ad-hoc fashion, indicating the lack of a targeted approach. These results highlight the need for strengthening cross-disciplinary connections between basic science and clinical studies, and the need for structured development and testing of therapeutic approaches to find more effective treatment interventions.

## Introduction

With several recent Phase 3 clinical trials of motor rehabilitation for stroke showing no differences between the treatment and the control group^1–3^, there is an urgent need to find more effective therapeutic interventions. A major challenge that has been highlighted in several recent commentaries^4–10^ is the need for interventions that are based on a targeted approach - i.e., interventions with a strong scientific premise and appropriate biological targets. Consistent with this observation, a recent review^11^ that examined approximately 194 randomized controlled trials from 1979 to 2013 found that only about 31% of the trials described a clear rationale, and that the inclusion of a rationale was associated with increased odds of a positive finding.

The purpose of this review was to more closely examine the basis for current therapeutic approaches in stroke rehabilitation. We made two major changes to the approach adopted by the previous review^11^ - (i) we focused on recent articles in the last 2 years (2019-2020) to reflect more contemporary approaches to stroke rehabilitation, and (ii) in addition to a quantitative description (i.e., describing if a particular study mentioned a specific rationale), we used a thematic analysis to provide more fine-grained analysis of how therapies are designed and evaluated in studies. Specifically, we focused on three issues - (i) the scientific rationale for the therapeutic intervention, (ii) the active ingredients of the therapeutic intervention, and (iii) the characteristics of different therapeutic interventions and the link between different therapeutic interventions in multimodal approaches.

## Methods

### Study selection

Our selection process is described in Figure 1. We extracted studies published in a time span of 2 years (1 Jan 2019-31 Dec 2020) from PubMed with the keywords listed in Figure 1. From the initial list of 539 studies, we used the following inclusion criteria: (i) prospective studies that randomly assigned participants to groups, (ii) the presence of at least one “control” or “usual care” group, (iii) outcomes that focused on motor function, and (iv) primary research studies that described the intervention (i.e., not follow-up of prior intervention studies). Exclusion criteria were: (i) the use of a drug/pharmacological intervention, and (ii) the use of alternative/complementary therapies.

**Figure 1.**
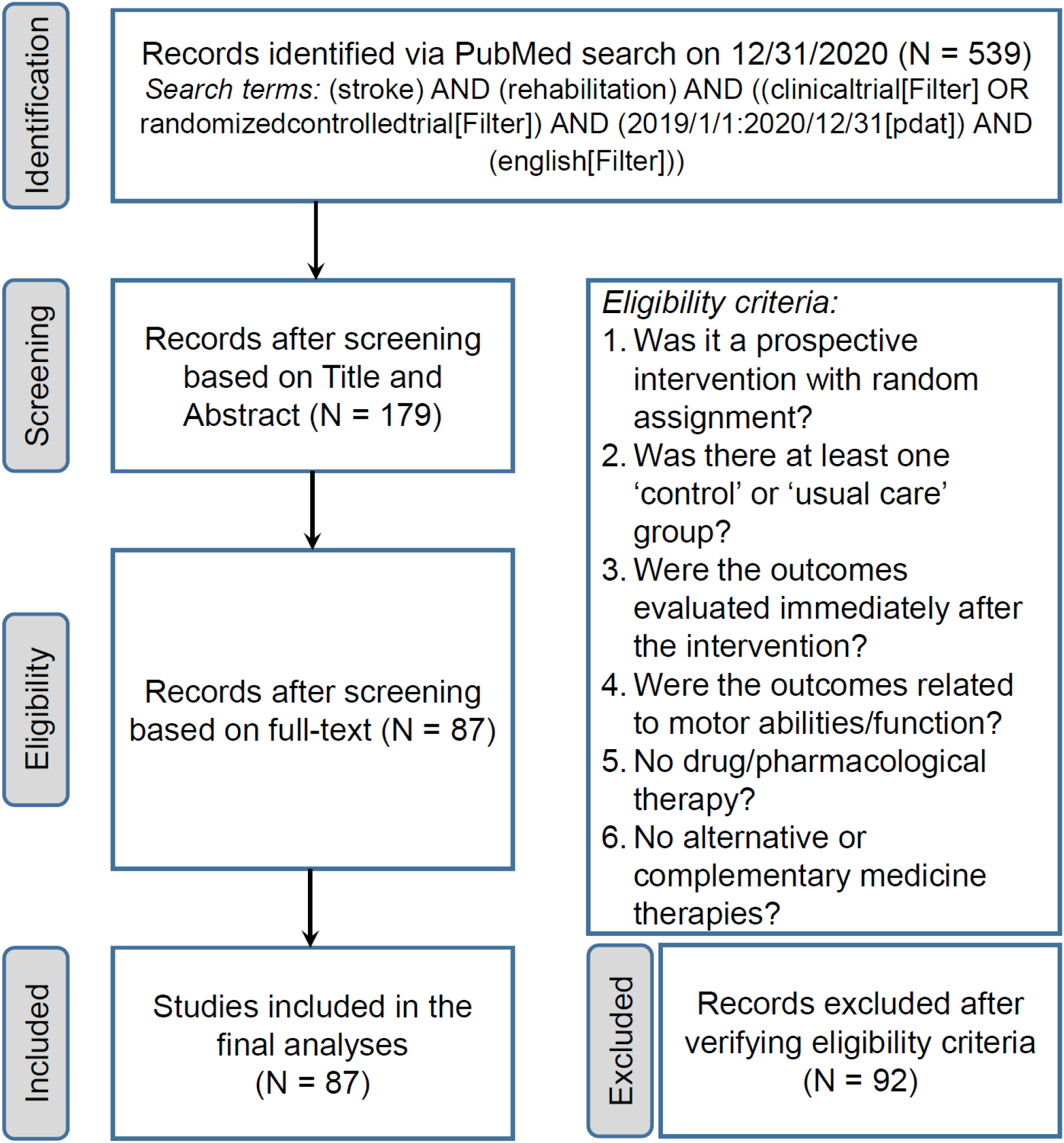
Flowchart of studies reviewed. Out of the 539 records retrieved from our original search, we used a total of 87 studies in the final analyses.

## Data analysis

### Descriptive parameters

We extracted the following parameters from each study - (i) demographic characteristics (age, time since stroke, and baseline motor function scores), (ii) intervention characteristics (upper or lower limb intervention, type of therapeutic intervention, and whether the intervention was multimodal), and (iii) study characteristics (sample size, and whether the result found a significant difference between the treatment and the control group in the primary outcome).

### Thematic analysis

#### Scientific *rationale*

We examined if the paper provided an explicit scientific rationale for why the particular treatment would work. We used two criteria for deciding if there was an explicit scientific rationale: (i) there was a specific hypothesis or prediction sentence about the group comparison, and (ii) there was an associated scientific basis related to this hypothesis or prediction. When we judged a study to have an explicit scientific rationale, we also examined if there was mention of ‘basic science’ studies in the Introduction section-either animal studies or motor learning studies in unimpaired humans. We focused on these two categories specifically to address the issue of how knowledge from these fields impacted the rationale for stroke rehabilitation.

#### Active Ingredients

We generated a list of nine active ingredients commonly used in neurorehabilitation (Table 1). For each therapeutic intervention, we listed the active ingredients that were mentioned in the Introduction. In addition, we also examined how many of these active ingredients mentioned were measured in the paper either in the Methods or by using any specific dependent variable. For example, if terms such as the intensity of practice was mentioned in the Introduction, we classified it as the active ingredient “dose” and we also examined whether the intervention mentioned the number of repetitions or the time duration of therapy. Similarly, if the paper mentioned greater engagement using their therapy, we classified this under the “motivation” ingredient and examined if their dependent variables measured if participants indeed had greater engagement. Data visualization was performed using the ggplot2 package^12^.

**Table 1.** List of the most common active ingredients used in therapeutic interventions.

| **Active Ingredient** | **“Rehab needs to...”** | **Examples** |
| --- | --- | --- |
| Dose | Be more intense | Intensity/time of therapy |
| Motivation | Be more fun and engaging | Gaming, virtual reality |
| Timing | Be done at the right time | Moving to acute phase |
| Real-world | Be integrated into the real-world | Home-based/ telerehabilitation |
| Movement | Elicit correct movements | Robotics |
| Feedback | Provide the right information | Mirror therapy |
| Neuroplasticity | Activate the right neural pathways | Brain stimulation |
| Task-oriented | Increase functional ability | Treadmill training |
| Fitness | Increase movement capacity | Strength training |

#### Therapies

Based on our search results, we examined 11 types of therapeutic interventions – robotics, virtual reality (VR), mirror therapy, non-invasive brain stimulation (NIBS), electromechanical stimulation (EMS), bilateral training (Bilateral), functional therapy (function), fitness training (Fitness), sensory cueing (Cue), action observation therapy (AOT), and other. If a paper had a multimodal intervention (i.e., it used more than 1 type of therapeutic intervention), we examined the two main interventions in the intervention.

## Results

### Descriptive characteristics

The average age of participants across studies was 60 ± 5 years. There was a wide variation in terms of time after stroke with about 45% of studies focusing on the ‘pre-chronic’ stage (i.e., prior to 6 months after stroke) (Figure 2A). Similarly, there was also a wide variation in terms of impairment as measured by the Fugl-Meyer score in studies involving the upper extremity, and gait speed in studies involving the lower extremity (Figure 2B-C). Most studies involved individuals with moderate impairments.

**Figure 2.**
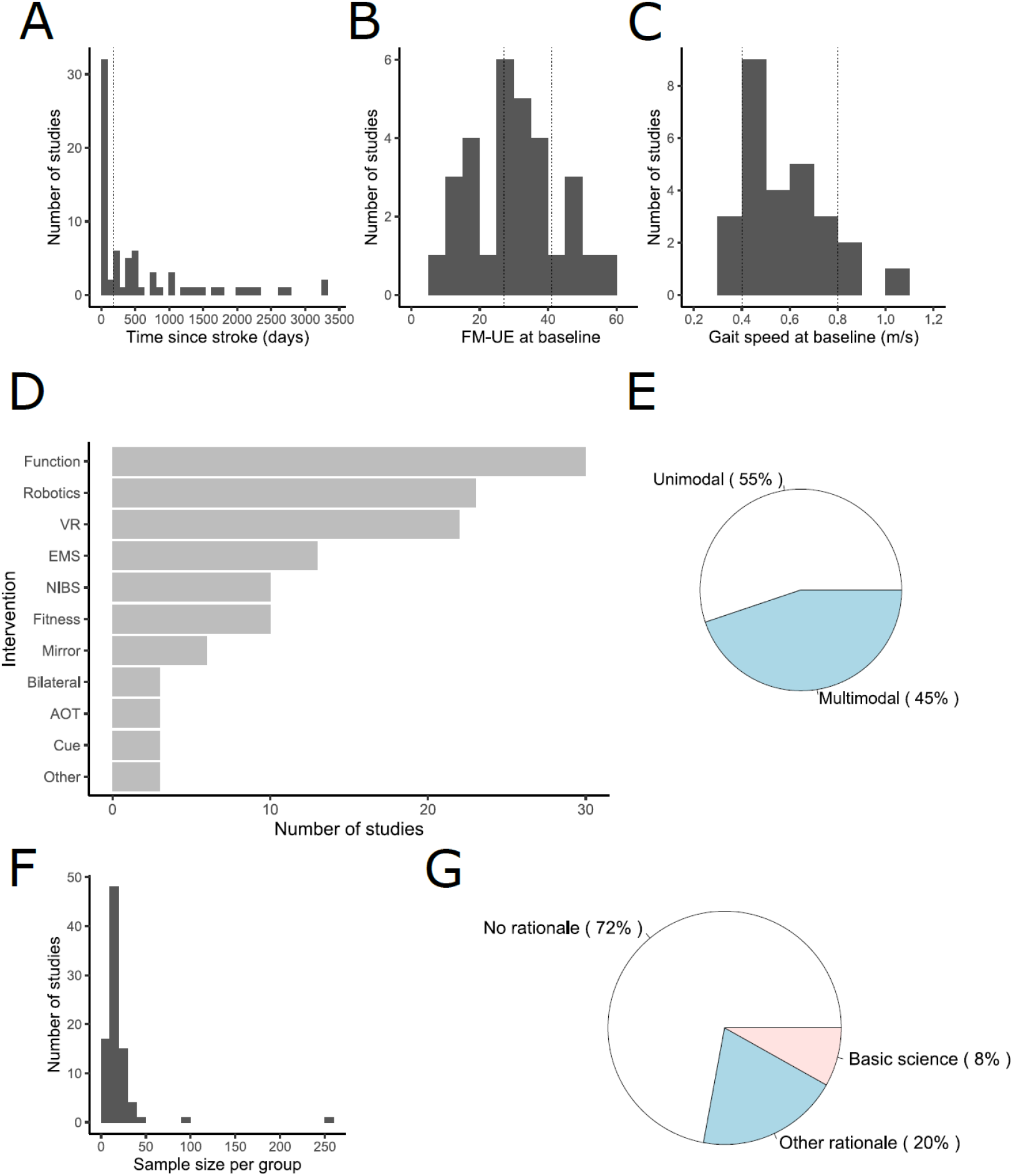
Sample, intervention, and study characteristics. (A) Histogram of the time after stroke at which participants were enrolled at baseline. About 45% of studies were to the left of the vertical dotted line which indicates the 6-month period after which the stroke is classified as being in the ‘chronic’ stage. (B) Fugl-Meyer upper extremity (FM-UE) score and (C) Gait speed of participants at baseline. Dotted lines in both panels indicate the region where the impairment is considered ‘moderate’. (D) Histogram of the different types of interventions used. The total number exceeds the total number of studies because several studies had more than one type of intervention. (E) Distribution of unimodal and multimodal interventions. (F) Average sample size per group. (G) Distribution of studies based on the scientific rationale provided. A basic science rationale here was defined as being related to animal studies or human motor learning. NIBS = noninvasive brain stimulation; EMS = electromechanical stimulation; VR = virtual reality; Mirror = mirror therapy; Bilateral = bilateral training; Function = functional therapy; Fitness = fitness training; Cue = sensory cueing; AOT = action observation therapy.

In terms of intervention characteristics, interventions tended to either focus exclusively on the upper (46%) or lower limb interventions (45%), with only a small number of studies focusing on the trunk or a combination (9%). The most common interventions were related to function (35%), robotics (26%), and virtual reality (25%) (Figure 2D). There was a relatively even split between studies that used multimodal therapy (45%) and those that focused on a single mode of therapy (55%) (Figure 2E).

Finally, in terms of study characteristics, we noticed that sample sizes were generally small (Median = 16, IQR = 9.5) (Figure 2F). However, despite the small sample size, a ‘positive’ result (i.e., the treatment group outperformed the control group on the primary outcome) was reported in 61% of studies.

### Thematic analysis

#### Scientific rationale

Only 28% of studies mentioned an explicit scientific rationale for the therapeutic intervention. 8% mentioned ‘basic science’ studies in their rationale (either animal or motor learning studies) and the remaining 20% used other reasons (mostly prior work related to stroke rehabilitation) in their rationale (Figure 2G).

#### Active ingredients

Out of the identified list of ingredients, the most frequently mentioned ingredients were neuroplasticity (38%) and dose (33%), while the least frequently mentioned ingredients were timing (7%) and fitness (14%).

Most studies on average mentioned 2 ingredients, while they measured only 1 ingredient. When we examined the number of times an ingredient was mentioned versus the number of it was actually measured, there was a discrepancy between the two quantities (Figure 3A). A bigger discrepancy between the two quantities can be indicative of evidence that an ingredient is frequently mentioned as an important component of rehabilitation but not tested directly. In this regard, the biggest discrepancies were found for the neuroplasticity and motivation ingredients (Figure 3B).

**Figure 3.**
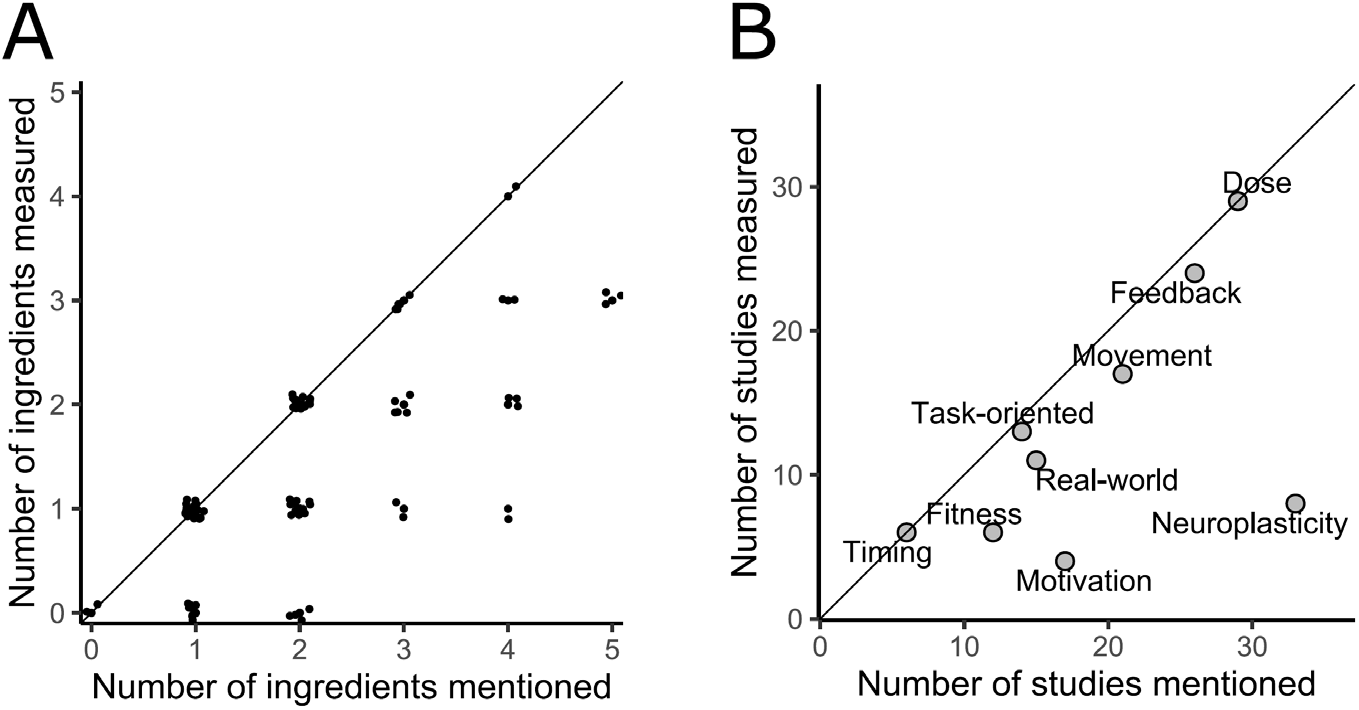
Analysis of active ingredients. **(A)** Number of active ingredients measured in the study vs. the number of ingredients mentioned in the Introduction section of the study. The diagonal line indicates the ‘identity’ line where the number of active ingredients measured is the same as the number of active ingredients mentioned. The fact that several studies are below this line indicates that there are ingredients that are mentioned in the Introduction but not measured. (B) Number of studies where a specific active ingredient was measured vs. mentioned in the Introduction. Ingredients like neuroplasticity and motivation that are significantly below the diagonal line indicate that they are frequently mentioned but rarely measured.

#### Therapies

The active ingredients for different therapeutic interventions are shown in Figure 4. Although no intervention was exclusively associated with a single active ingredient, we found that the interventions varied a lot in terms of their specificity. For example, mirror therapy and NIBS were highly specific with most studies using these therapies mentioning mostly on one or two active ingredients, whereas function and electrical stimulation were at the other extreme with several active ingredients being mentioned across studies.

**Figure 4.**
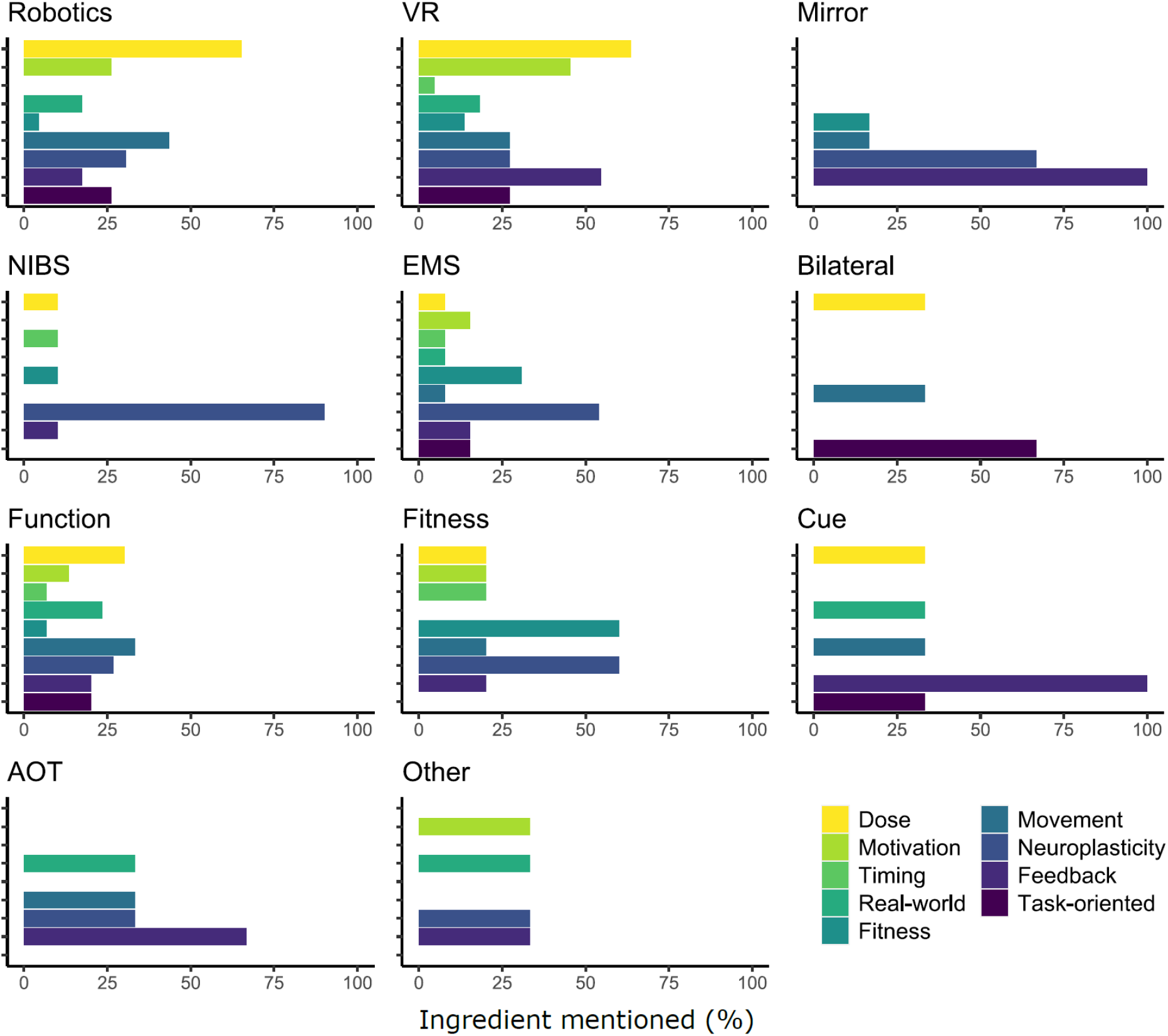
Ingredient analysis of different interventions. Within each plot, each bar represents a different active ingredient, and the length of the bar represents the percentage of studies using that therapy where that ingredient was mentioned. Some interventions have very high specificity in terms of the ingredients (e.g., NIBS and Mirror therapy) as indicated by a few dominant ingredients, while others have very low specificity (e.g., EMS and Function). NIBS = noninvasive brain stimulation; EMS = electromechanical stimulation; VR = virtual reality; Mirror = mirror therapy; bilateral = Bilateral training; Function = functional therapy; Fitness = fitness training; Cue = sensory cueing; AOT = action observation therapy.

When we examined the connection between the different therapies used in multimodal therapies (Figure 5), the largest interconnections were found between robotics and virtual reality (6 studies), and robotics and function (10 studies). However, there were almost links between all possible pairs of therapies, indicating that these combinations tend to be combined in an ad-hoc manner.

**Figure 5.**
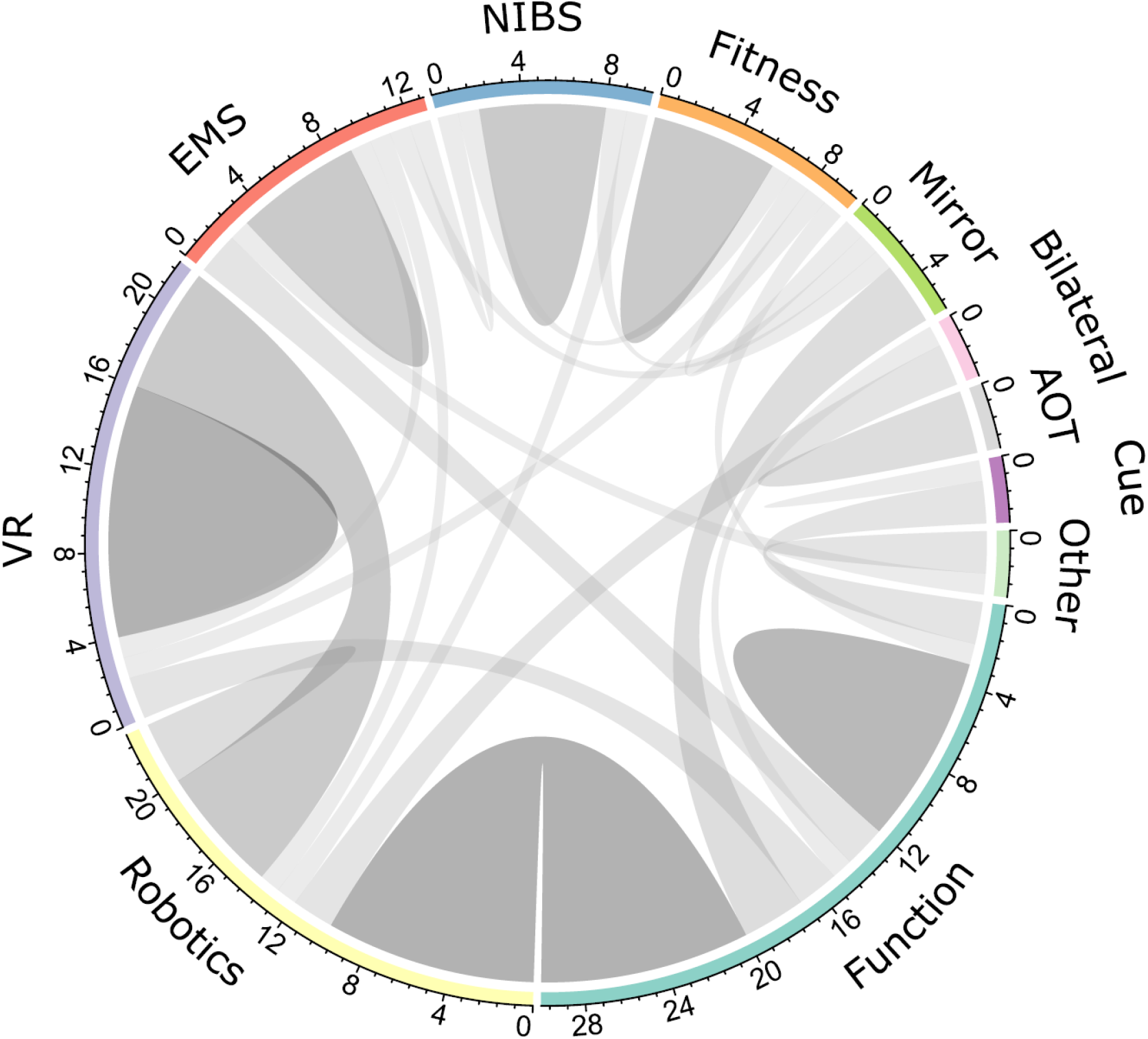
Chord plot showing the interconnection between the different therapeutic interventions. The perimeter of the circle is color coded by the different therapeutic interventions, with the length proportional to the number of studies using that therapy. Chords connecting two different therapies indicate the number of studies that uses a combination of both therapies. Unimodal therapies (i.e., studies that only use one type of therapy) are indicated by a self-directed link. Overall, the existence of links between almost all possible pairs of therapies indicates that these combinations tend to be combined in an ad-hoc manner. This plot was generated using the circlize package in R^31^. NIBS = noninvasive brain stimulation; EMS = electromechanical stimulation; VR = virtual reality; Mirror = mirror therapy; Bilateral = bilateral training; Function = functional therapy; Fitness = fitness training; Cue = sensory cueing; AOT = action observation therapy.

## Discussion

The purpose of this review was to use a thematic analysis to examine the scientific rationale behind current therapeutic interventions, the active ingredients in these interventions, and the link between different therapeutic interventions. Our results showed three major findings - (i) a majority of studies did not provide an explicit rationale for why the treatment would be expected to work, (ii) although several active ingredients were mentioned for each therapy, there was not a close correspondence between the active ingredients mentioned versus those reported or measured in the study, and (iii) multimodal approaches that involved more than one therapeutic intervention tended to be combined in an ad-hoc manner, indicating the lack of a targeted approach. These are further discussed below.

### Missing Rationale

Consistent with prior work ^11^, our analysis indicated that studies often did not include an explicit rationale with a prediction of what their intervention was addressing. The rationale for the majority of studies highlighted a limitation of a prior study and/or the innovation of the current study but did not provide the full rationale for why the current intervention should be expected to work. Moreover we found, perhaps surprisingly, that there was little mention of links to ‘basic science’ studies (either animal or human motor learning studies), which is considered a key-feature especially in early phase trials^10^. This may reflect a bias in terms of citing some types of research, but also potentially indicates a breakdown in translating evidence from basic science studies to stroke rehabilitation.

### Active Ingredients: Walking the talk?

When examining active ingredients, there were usually multiple active ingredients associated with each therapy. Depending on the type of therapy, these often ranged up to 5 active ingredients, which indicates that many studies tend to ‘cover all bases’ when justifying the intervention. This analysis was also evident in the analysis of the individual interventions where most therapies were associated with multiple active ingredients. Our view is that therapeutic interventions (at least in early phase studies) should have greater specificity in terms of the ingredients because greater specificity increases the chance that any differences in overall outcome can be attributed to a specific ingredient, which in turn opens the door for designing new therapies for larger trials.

Multimodal therapies tended to be even more mixed in terms of active ingredients, but with no specific patterns in terms of combining therapies. The ideal multimodal approach of two interventions A and B is one where there is a “synergistic” effect - i.e., the combined effect of A+B is greater than what can be obtained through A or B alone. However, identifying such combinations requires clear specification of the unique active ingredients of each therapy, and a rationale for why the combination of these two ingredients is critical for effective rehabilitation. The lack of such specific patterns once again highlights the lack of specificity in terms of the active ingredients of each therapy.

In addition, we also found poor correspondence between ‘mentioned ingredients’ and ‘measured ingredients’. This situation leads to the creation of “buzzword ingredients’’ - i.e., ingredients that are mentioned frequently, but where evidence is not accumulated towards testing their effect. For example, studies that mentioned neuroplasticity as an active ingredient of the therapy frequently did not use any related direct neural measures of neuroplasticity (such as EEG, fMRI or TMS). Although neuroplasticity plays a critical role in recovery in some contexts, mentioning it as a general ingredient in all therapeutic interventions (or alternatively to use the term as a general phrase to indicate any type of recovery or learning) decreases the chances of understanding what effects are actually due to neuroplasticity.

### Recommendations for future research

In light of these findings, we suggest three recommendations for future work in order to facilitate the search for effective therapeutic interventions.

### Need for better dialog between basic science and clinical interventions

The lack of rationale highlights the need for a tighter link between rationale mentioned and the actual intervention delivered. Although stroke rehabilitation is a complex process where understanding the role of different ingredients may not always be direct, attempting to identify and uncover the effects of these ingredients is critical from a scientific perspective. For example, there have been controversies even with some of the most well-established active ingredients like the dosage of practice^13–16^.This is partly because even when the evidence from studies in animal models is robust, translating the work from animal models into humans is non-trivial and requires careful attention to many factors^7^.

Another important piece in the translational pipeline is motor learning studies in unimpaired humans. These studies are especially critical for certain active ingredients (such as motivation or feedback), where reliance on animal models may not be optimal^17^. However, while there is agreement that “principles of motor learning” form a basis for effective neurorehabilitation practice^18–22^, there tends to be a large gulf between experiments in motor learning (which are largely focused on single-day experiments with well-controlled laboratory tasks) and real-world stroke rehabilitation. Therefore, while it is important that clinical researchers increase their awareness of motor learning research, it is also equally critical for motor learning researchers to bridge this gap in experimental paradigms for more effective translational research.

### Measurement and evaluation of the effect of active ingredients

Once active ingredients of a specific intervention are identified, there should be close alignment with direct measurement of the effects of these active ingredients. Similar to the need for ‘modality-specific’ outcome measures^23^ (for e.g., using motor-specific outcomes for motor rehabilitation), outcome measures should also be ‘ingredient-specific’ and directly relate to the active ingredient proposed. Depending on the type of active ingredient, these may include measurements both ‘during’ the intervention (e.g., to evaluate whether training with VR increases motivation) or at the ‘end’ of the intervention (e.g., to evaluate whether an intervention causes neural plasticity). By tracking these ingredient-specific measures separately from the overall outcome of the intervention, different decisions can be made from both trials where the intervention ‘worked’ and those that did not, as highlighted in Table 2.

**Table 2.**
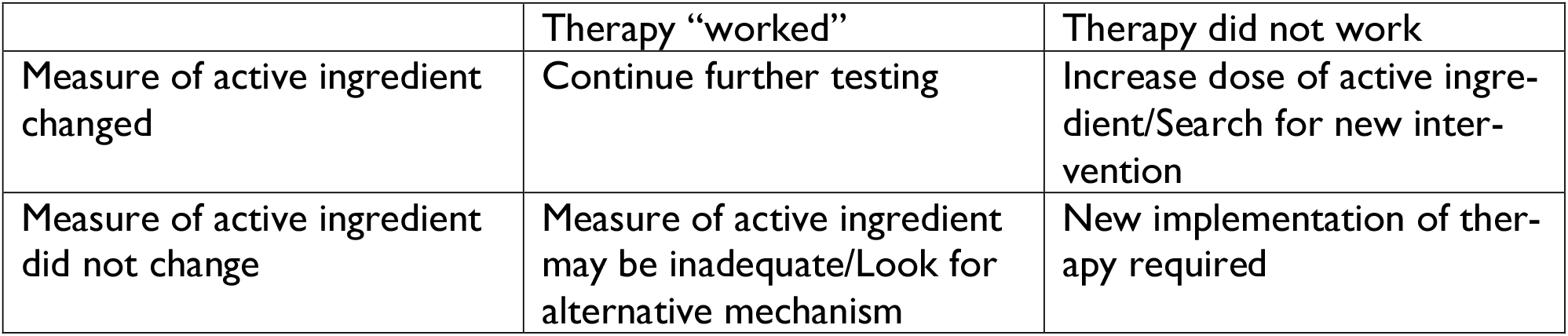
The decision matrix for subsequent course of action depends on two factors – whether the therapy ‘worked’ (i.e., in terms of the outcome), and whether the measure of the active ingredient changed.

|  | Therapy “worked” | Therapy did not work |
| --- | --- | --- |
| Measure of active ingredient changed | Continue further testing | Increase dose of active ingredient/Search for new intervention |
| Measure of active ingredient did not change | Measure of active ingredient may be inadequate/Look for alternative mechanism | New implementation of therapy required |

Table 2 also highlights an important aspect of why measurement and evaluation of active ingredients is critical. Rather than serve as “post-hoc” explanations of findings, addressing these issues in early-phase pre-clinical trial stage can help narrow down the optimal decision at the end of a trial^24^. For example, if the active ingredient changed but there was no effect on overall outcome, there are two potential decisions to be made - (i) continue with the therapy, but increase the dosage of the ingredient, or (ii) switch to a different therapy. This decision can be made more effectively if there are already prior dose-response studies indicating whether the dosage of the active ingredient was sufficient to induce any change in motor function.

It is also worth noting two points that we did not consider in our current review. First, measuring the effects of some types of active ingredients require more than simply adding ingredient-specific dependent variables. Instead, assessing these active ingredients may require fundamental changes to the experimental design itself, such as the addition of separate control groups. For example, a trial focusing on ‘timing’ as an active ingredient (i.e., starting rehabilitation in the acute or subacute phases of stroke) may not only need a measure of the time when rehabilitation was started, but also need appropriate control groups to measure the effect of timing – i.e., separating spontaneous biological recovery from the effects due to the early intervention. Second, greater specificity when reporting ingredients is also critical – for example, while most studies reported the dosage of the intervention in terms of the overall number of sessions and the time per session, ‘dose’ is a multidimensional construct which requires information about parameters such as session density, task difficulty and intensity ^25^. Therefore, developing guidelines and reporting standards for the active ingredients is an important step to better understand their effects on recovery.

### Improving overall general study quality

Finally, our review indicated that the study population used in these studies was characterized by small sample sizes and people with mild-to-moderate impairments. These highlight two potential issues - first, many rehabilitation interventions currently address a small fraction of the stroke population. For example, the recruitment rate for most large Phase 3 clinical trials tends to be smaller than 10% of the screened population ^2,3,26^. While it is inevitable that some types of therapies require a certain degree of motor function, this also suggests the need to find more therapies that are applicable to a wider population. Second, the small recruitment also raises the issue of sample size, with a median of 16/group in our review, with ∼75% of studies being under 20. For comparison, in the VA ROBOTICS trial^1^, the comparison between the experimental group and the usual care group (powered to detect a large effect size of Cohen’s d of 1 at 90% power), required a sample size of 23/group. Although some of these may be mitigated by using specific strategies such as stratification at baseline, the sample sizes needed for most studies typically exceed these numbers, sometimes by an order of magnitude^27^. As mentioned in several commentaries^28,29^, low sample size not only decreases statistical power but also increases the risk of false positives. Therefore, even though the estimate in review of positive trials is around 60%, the inflation in false positive rate due to the small sample sizes make it unclear as to what proportion of these trials have a true positive effect. This points to the need for larger sample sizes even in early phase trials for determining which therapeutic interventions might have the most promise in large scale trials.

In conclusion, our review highlights several factors that currently limit the ability to find effective treatments for stroke rehabilitation. While it is important to acknowledge the barriers and challenges associated with running clinical trials in neurorehabilitation^30^, we believe that the issues posed here further highlight the need to coordinate resources, design and run more informative studies, which will eventually pave the way for a more optimal search for effective interventions.

## Data Availability

All data related to this article has been posted as Supplementary material

## Acknowledgment

This material is based upon work partly supported by the National Science Foundation under Grants No. 1823889 (RR), 1804053 (CK), and by the National Institutes of Health R01EB019834 (CK).

## Supplementary information

The list of articles included in the review along with the data is provided at https://osf.io/vg46a/

## References

1. Lo AC, Guarino PD, Richards LG, et al. Robot-Assisted Therapy for Long-Term Upper-Limb Impairment after Stroke. N Engl J Med. 2010;362(19):1772–1783. doi:10.1056/NEJMoa0911341

2. Duncan PW, Sullivan KJ, Behrman AL, et al. Body-Weight–Supported Treadmill Rehabilitation after Stroke. N Engl J Med. 2011;364(21):2026–2036. doi:10.1056/NEJMoa1010790

3. Winstein CJ, Wolf SL, Dromerick AW, et al. Effect of a Task-Oriented Rehabilitation Program on Upper Extremity Recovery Following Motor Stroke: The ICARE Randomized Clinical Trial. JAMA. 2016;315(6):571–581. doi:10.1001/jama.2016.0276

4. Stinear CM, Lang CE, Zeiler S, Byblow WD. Advances and challenges in stroke rehabilitation. Lancet Neurol. 2020;19(4):348–360. doi:10.1016/S1474-4422(19)30415-6

5. Ward NS, Carmichael ST. Blowing up Neural Repair for Stroke Recovery: Preclinical and Clinical Trial Considerations. Stroke. 2020;51(10):3169–3173. doi:10.1161/STROKEAHA.120.030486

6. Bowden MG, Woodbury ML, Duncan PW. Promoting neuroplasticity and recovery after stroke: future directions for rehabilitation clinical trials. Curr Opin Neurol. 2013;26(1):37–42. doi:10.1097/WCO.0b013e32835c5ba0

7. Krakauer JW, Carmichael ST, Corbett D, Wittenberg GF. Getting neurorehabilitation right: what can be learned from animal models? Neurorehabil Neural Repair. 2012;26(8):923–931. doi:10.1177/1545968312440745

8. Krakauer JW, Carmichael ST. Broken Movement: The Neurobiology of Motor Recovery after Stroke. MIT Press; 2017.

9. Ward NS. Restoring brain function after stroke — bridging the gap between animals and humans. Nat Rev Neurol. 2017;13(4):244–255. doi:10.1038/nrneurol.2017.34

10. Tsay JS, Winstein CJ. Five Features to Look for in Early-Phase Clinical Intervention Studies. Neurorehabil Neural Repair. 2021;35(1):3–9. doi:10.1177/1545968320975439

11. Borschmann K, Hayward KS, Raffelt A, Churilov L, Kramer S, Bernhardt J. Rationale for Intervention and Dose Is Lacking in Stroke Recovery Trials: A Systematic Review. Stroke Res Treat. 2018;2018:e8087372. doi:10.1155/2018/8087372

12. Wickham H. Ggplot2: Elegant Graphics for Data Analysis. 2nd ed. Springer International Publishing; 2016. doi:10.1007/978-3-319-24277-4

13. Lohse KR, Lang CE, Boyd LA. Is more better? Using metadata to explore dose-response relationships in stroke rehabilitation. Stroke. 2014;45(7):2053–2058. doi:10.1161/STROKEAHA.114.004695

14. Lang CE, Strube MJ, Bland MD, et al. Dose response of task-specific upper limb training in people at least 6 months poststroke: A phase II, single-blind, randomized, controlled trial. Ann Neurol. 2016;80(3):342–354. doi:10.1002/ana.24734

15. Daly JJ, McCabe JP, Holcomb J, Monkiewicz M, Gansen J, Pundik S. Long-Dose Intensive Therapy Is Necessary for Strong, Clinically Significant, Upper Limb Functional Gains and Retained Gains in Severe/Moderate Chronic Stroke. Neurorehabil Neural Repair. 2019;33(7):523–537. doi:10.1177/1545968319846120

16. Ward NS, Brander F, Kelly K. Intensive upper limb neurorehabilitation in chronic stroke: outcomes from the Queen Square programme. J Neurol Neurosurg Psychiatry. 2019;90(5):498–506. doi:10.1136/jnnp-2018-319954

17. Winstein C, Varghese R. Been there, done that, so what’s next for arm and hand rehabilitation in stroke? NeuroRehabilitation. 2018;43(1):3–18. doi:10.3233/NRE-172412

18. Krakauer JW. Motor learning: its relevance to stroke recovery and neurorehabilitation. Curr Opin Neurol. 2006;19(1):84. doi:10.1097/01.wco.0000200544.29915.cc

19. Kitago T, Krakauer JW. Motor learning principles for neurorehabilitation. Handb Clin Neurol. 2013;110:93–103. doi:10.1016/B978-0-444-52901-5.00008-3

20. Fisher BE, Morton SM, Lang CE. From Motor Learning to Physical Therapy and Back Again: The State of the Art and Science of Motor Learning Rehabilitation Research. J Neurol Phys Ther JNPT. 2014;38(3):149–150. doi:10.1097/NPT.0000000000000043

21. Muratori LM, Lamberg EM, Quinn L, Duff SV. Applying principles of motor learning and control to upper extremity rehabilitation. J Hand Ther. 2013;26(2):94–103. doi:10.1016/j.jht.2012.12.007

22. Winstein C, Lewthwaite R, Blanton SR, Wolf LB, Wishart L. Infusing motor learning research into neurorehabilitation practice: a historical perspective with case exemplar from the accelerated skill acquisition program. J Neurol Phys Ther JNPT. 2014;38(3):190–200. doi:10.1097/NPT.0000000000000046

23. Cramer SC, Koroshetz WJ, Finklestein SP. The Case for Modality-Specific Outcome Measures in Clinical Trials of Stroke Recovery-Promoting Agents. Stroke. 2007;38(4):1393–1395. doi:10.1161/01.STR.0000260087.67462.80

24. Dalton EJ, Churilov L, Lannin NA, Corbett D, Campbell BCV, Hayward KS. Multidimensional Phase I Dose Ranging Trials for Stroke Recovery Interventions: Key Challenges and How to Address Them. Neurorehabil Neural Repair. Published online June 4, 2021:15459683211019362. doi:10.1177/15459683211019362

25. Hayward KS, Churilov L, Dalton EJ, et al. Advancing Stroke Recovery Through Improved Articulation of Nonpharmacological Intervention Dose. Stroke. 2021;52(2):761–769. doi:10.1161/STROKEAHA.120.032496

26. Wolf SL, Winstein CJ, Miller JP, et al. Effect of constraint-induced movement therapy on upper extremity function 3 to 9 months after stroke: the EXCITE randomized clinical trial. JAMA. 2006;296(17):2095–2104. doi:10.1001/jama.296.17.2095

27. Winters C, Heymans MW, EEH van Wegen, Kwakkel G. How to design clinical rehabilitation trials for the upper paretic limb early post stroke? Trials. 2016;17(1):468. doi:10.1186/s13063-016-1592-x

28. Button KS, Ioannidis JPA, Mokrysz C, et al. Power failure: why small sample size undermines the reliability of neuroscience. Nat Rev Neurosci. 2013;14(5):365–376. doi:10.1038/nrn3475

29. Ioannidis JPA. Why most published research findings are false. PLoS Med. 2005;2(8):e124. doi:10.1371/journal.pmed.0020124

30. Wolf SL, Winstein CJ, Miller JP, Blanton S, Clark PC, Nichols-Larsen D. Looking in the rear view mirror when conversing with back seat drivers: the EXCITE trial revisited. Neurorehabil Neural Repair. 2007;21(5):379–387. doi:10.1177/1545968307306238

31. Gu Z, Gu L, Eils R, Schlesner M, Brors B. circlize implements and enhances circular visualization in R. Bioinformatics. 2014;30(19):2811–2812. doi:10.1093/bioinformatics/btu393

